# Single Ventricle Reconstruction III: Brain Connectome and Neurodevelopmental Outcomes: Design, Recruitment, and Technical Challenges of a Multicenter, Observational Neuroimaging Study

**DOI:** 10.1101/2023.04.12.23288433

**Authors:** Vanessa Schmithorst, Rafael Ceschin, Vince Lee, Julia Wallace, Aurelia Sahel, Thomas Chenevert, Hemant Parmar, Jeffrey I. Berman, Arastoo Vossough, Deqiang Qiu, Nadja Kadom, Patricia Ellen Grant, Borjan Gagoski, Peter LaViolette, Mohit Maheshwari, Lynn A. Sleeper, David Bellinger, Dawn Ilardi, Sharon O’Neil, Thomas A. Miller, Jon Detterich, Kevin D. Hill, Andrew M. Atz, Marc Richmond, James Cnota, William T.Mahle, Nancy Ghanayem, William Gaynor, Caren S. Goldberg, Jane W. Newburger, Ashok Panigrahy, the Pediatric Heart Network SVRIII Brain Connectome Study Investigators

**Author notes:** **Corresponding Author:** Ashok Panigrahy, M.D., Department of Pediatric Radiology, Children’s Hospital of Pittsburgh of UPMC, 45^th^ Street and Penn Avenue, Pittsburgh, PA 15201 USA.

## Abstract

Patients with hypoplastic left heart syndrome who have been palliated with the Fontan procedure are at risk for adverse neurodevelopmental outcomes, lower quality of life, and reduced employability. We describe the methods (including quality assurance and quality control protocols) and challenges of a multi-center observational ancillary study, SVRIII (Single Ventricle Reconstruction Trial) Brain Connectome. Our original goal was to obtain advanced neuroimaging (Diffusion Tensor Imaging and Resting-BOLD) in 140 SVR III participants and 100 healthy controls for brain connectome analyses. Linear regression and mediation statistical methods will be used to analyze associations of brain connectome measures with neurocognitive measures and clinical risk factors. Initial recruitment challenges occurred related to difficulties with: 1) coordinating brain MRI for participants already undergoing extensive testing in the parent study, and 2) recruiting healthy control subjects. The COVID-19 pandemic negatively affected enrollment late in the study. Enrollment challenges were addressed by 1) adding additional study sites, 2) increasing the frequency of meetings with site coordinators and 3) developing additional healthy control recruitment strategies, including using research registries and advertising the study to community-based groups. Technical challenges that emerged early in the study were related to the acquisition, harmonization, and transfer of neuroimages. These hurdles were successfully overcome with protocol modifications and frequent site visits that involved human and synthetic phantoms.

**Trial registration number:** **ClinicalTrials.gov Registration Number: NCT02692443**

## Introduction

In the current era of cardiothoracic surgery, children with hypoplastic left heart syndrome (HLHS) are more likely to survive into adulthood than in previous eras. Improved survival has unmasked significant morbidity, including neurodevelopmental and psychosocial impairments that have been shown to affect school performance, employment, and quality of life.[1-10] The etiology of neurodevelopmental impairment in single ventricle patients is multifactorial and includes reduced *in-utero* blood flow, low birth weight, presence of genetic abnormalities, prolonged cyanosis, congestive heart failure, unstable hemodynamics during the perioperative period, and need for multiple cardiac catheterization and serial operations with prolonged hospital stays. [11-16] Modeling of neurodevelopmental outcomes in single ventricle patients that incorporates these risk factors is only modestly predictive, explaining less than one-third of the variation in outcomes.[17,18] Many reports have documented widespread brain abnormalities in individuals with single ventricle throughout their lifespan, suggesting brain topology may be a potent biomarker that can predict neurodevelopmental outcome. [19-27] [28,29] However, few of these neuroimaging studies have linked brain MRI findings to neurodevelopmental outcomes[30,31] or to specific clinical factors, suggesting that new approaches for evaluating the brain in patients with complex CHD are needed.

The Single Ventricle Reconstruction (SVR) III Brain Connectome study aims to bridge this gap by taking full advantage of methodological and conceptual developments showing that the human brain is intrinsically organized into large-scale, coherent brain networks (topology) that can be understood using methods such as graph theory analysis.[32-38] This study utilizes a global, “systems-level” approach involving characterization of brain network connectivity or *brain “connectome*” in SVR III study participants.[32-35] We previously applied brain connectivity graph analysis to adolescent participants of the landmark Boston Circulatory Arrest Study (BCAS), in which neonates with dextro-transposition of the great arteries (d-TGA) were randomized to two different perfusion strategies and followed closely with serial neurodevelopmental assessments until the adolescent period. [39-42] Our findings in the BCAS cohort demonstrated that brain connectivity/graph analysis could distinguish multi-domain cognitive deficits from specific neurobehavioral phenotypes (e.g., ADHD), and also delineate specific relationships between neonatal peri-operative variables and long-term neurocognitive outcomes in adolescents with d-TGA. Other recent studies have described anomalous diffusion tensor-based connectome in CHD neonates and infants in both preoperative and postoperative periods, finding distinct patterns of structural network topology alterations. [43-47] Recent literature also suggests that genetic factors might impact the structural connectome in CHD.[45] However, there remains a dearth of brain connectomic analyses in pediatric/adolescent CHD. Therefore, we anticipate that applying brain connectivity analysis to the SVR cohort will lead to new insights into the understanding the relationships between clinical risk factors and cognitive/behavioral outcomes in children with HLHS and other related single right ventricle cardiac malformations.

Here we present the study design for the NHLBI-funded Pediatric Heart Network (PHN) SVRIII Brain Connectome study, which has finished enrollment and data collection. We discuss our experience with recruitment and technical challenges and our implementation of solutions that resulted in meeting our adjusted enrollment goals approximately one year after the completion of the parent study (during the Covid-19 pandemic). [48] We also present our plans for final data processing and final statistical analysis.

## Methods and Design

### Study design and funding

The NHLBI-funded PHN SVR III study, “Long-term Outcomes of Children with HLHS and the Impact of Norwood Shunt Type,**”** is a prospective follow-up study of an existing cohort of children with HLHS and other single RV anomalies who were enrolled as newborns in a randomized clinical trial of Norwood procedure with a modified Blalock-Taussig-Thomas shunt (MBTTS) *versus* a right ventricular-to-pulmonary artery shunt (RVPAS). The SVR III study was designed to determine whether the shunt assignment at the time of the Norwood operation is associated with cardiac function, transplant-free survival, exercise function, and neurodevelopmental outcomes at ages 10 to 12 years. Transplant-free survivors of the original SVR trial were invited to participate in the multidisciplinary evaluation including performance of a cardiac MRI, echocardiogram, exercise testing, and neurocognitive evaluations (Figure 1). In the ancillary Brain Connectome Study to the SVR III study, selected SVR sites added a brain imaging component to the standard follow-up testing in the parent SVR III study. Specifically, we compared findings in SVR participants using brain imaging in concert with a complete neurodevelopmental battery and clinical information collected through the SVR III study to findings in healthy controls.

**Figure 1:**
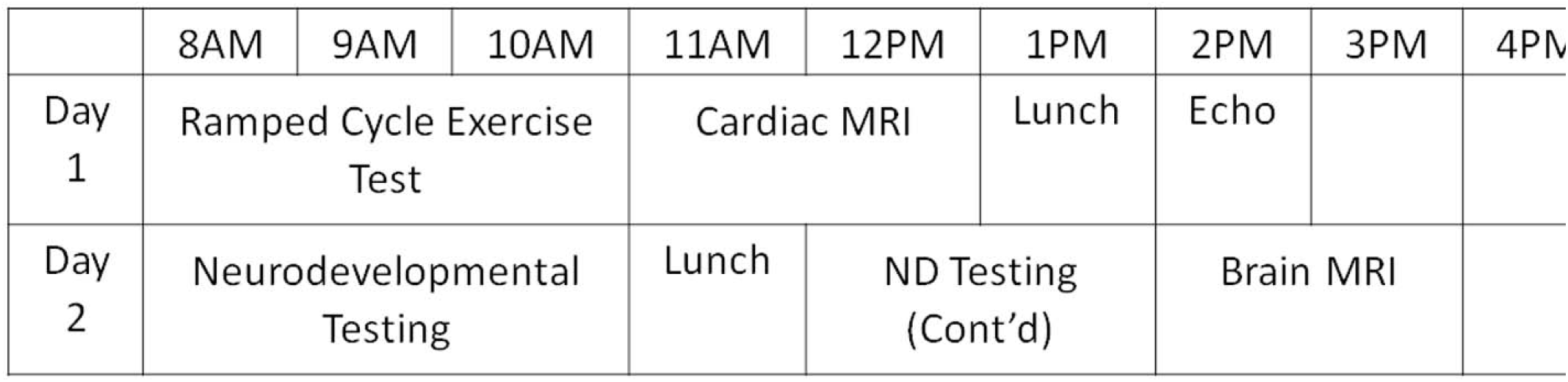
SVRIII Study Procedures: Schematic showing the parent SVRIII study procedures that were prioritized before ancillary study procedures (brain MRI)

We will pursue the mapping of brain connectivity using two state-of-the-art neuroimaging techniques: (1) *structural connectivity* using diffusion tensor imaging (DTI) to assess white matter tracts,[49-53] and (2) *functional connectivity* using large-amplitude spontaneous low-frequency (<0.1 Hz) fluctuations in the functional MRI or “resting” BOLD signal that is temporally correlated across functionally related regions of the brain.[54-58] These data will be analyzed using cutting edge quantitative complex network construction with graph theory to construct a brain connectome to characterize brain network topology. [32-35] By combining these innovative techniques within the setting of the PHN SVR long-term follow-up study (SVR III), our specific aims are: **1**. To characterize the global brain network topology of the SVR III cohort compare to healthy referents; **2**. To determine which neurocognitive and behavioral outcomes are predicted by global brain network topology; **3**. To determine which *patient factors* (e.g., birth weight, gestational age, and maternal education) and *medical factors* (e.g., intraoperative techniques during the Norwood procedure, hemodynamic complications, types and a number of interventions, and measures of global morbidity) predict global brain network topology; and, **4**. To precisely characterize inter-relationships between global brain network topology, patient/medical factors, and adverse neurocognitive/behavioral outcomes and quality of life.

### Screening, consent, and entry criteria for parent and ancillary study

The SVR III study subjects were initially eligible to participate in the Brain Connectome Study at 10 to 12.0 years of age. Over the course of the study period, the participation window was extended to age 12.5 years and then, ultimately, to any age reached by a participant by the close of the study in September 2020 when the oldest SVR subject was 15 years of age. The Principal Investigator at each site, their designees, and the study coordinator were responsible for participant recruitment into the ancillary study. All SVR III subjects were contacted to assess vital status. The transplant-free survivors were approached to participate in the ancillary study at in-person assessment SVR III study visits. SVR Trial subjects who underwent cardiac transplantation or biventricular conversion were excluded.

### Sites, Participants and Imaging Acquisition Protocol

PHN sites with more than ten eligible SVR III participants were initially invited for site participation in the SVR III Brain Connectome study. In addition, each site was asked to complete a detailed MRI questionnaire related to 3T neuro scanner capabilities (including vendor type, ability to run resting BOLD, 45 direction isotropic).

DTI, and research capacity for running a special customized multiband DTI -256 direction). We also queried for availability and interest of neuro-based personnel, including neuroradiologist, MR physicist, and MR technologists presence and capability. We developed an imaging protocol in which at least one of the connectome sequences at 3T (42 direction DTI) could be performed at all potential PHN recruiting sites. We also developed the rest of the neuroimaging protocol to facilitate the acquisition of multi-band-multi-shell HARDI diffusion imaging and also multiband resting state, in addition to volumetric T2 and T1 3D imaging, which was in alignment with the Adolescent Brain Cognitive Development (ABCD) study[59], accounting for the variability of the gradient strengths of the scanner (Supplemental Table 1).

### Multicenter MRI Quality Assurance and Quality Control (QA/QC)

For this study we used Siemens and Phillips 3T MRI systems. We conducted a methodological PHN inter-site reliability study to assure that the studies performed on these systems could be compared. [60,61] As proof of concept we analyzed human multimodal data (resting BOLD, DTI, and MR spectroscopy) from five PHN sites with standardized protocols for Siemens and Philips units on a small sample of control subjects age matched to the SVR subjects (n=10). The temporal signal-to-noise ratio (tSNR) of the Siemens and Philips resting BOLD data was comparable (Siemens: average SNR = 169; Philips: average tSNR = 161; 4 mm X 4 mm X 4 mm voxel size). We also found that the DTI data had a similar distribution of FA values for the two vendors. In addition, we noted that the quality of spectra showed no significant differences in line-width, SNR, or reliability of measurements suggesting the feasibility of high-quality multi-site MRS data. These data suggested that reliable neuroimaging data could be obtained across the PHN sites, and these metrics were integrated into our QA/QC protocol.

Proper QA/QC procedures are a complex multi-step process that involves both phantom and subject data. We adapted a QA procedure [62] used by multiple NIH-funded multicenter studies including the Human Brain Connectome, TRACK-TBI, Pediatric Brain Tumor Consortium (PBTC), and the ABCD study. [63-72] For our prospective QA plan, each site scanned at least two phantoms (ACR-anatomic and f-BIRN-functional) for QA purposes on months that a subject was scanned (approximately once/month/site). While our initial plan was to obtain diffusion QA data using the NIST Phantom, we utilized a synthetic HARDI phantom, given the single band and multiple band/multi-shell protocol that was incorporated into the study. [73-75] For the DTI phantom and human studies, we evaluated multiple values: (1) SNR at the center and periphery of the phantom; (2) comparison of image distortion in phase-encoding direction between EPI and spin echo image; (3) comparison of image distortion between nonzero b-value DWI and b=0 image caused by gradient encoding directions. From this, we corrected for image distortion in EPI readout caused by B_0_ inhomogeneity, distortion caused by eddy currents induced by diffusion-encoding gradients, uniformity of b-value along different diffusion-encoding directions, and correct calibration for accurate diffusivity measurements. For resting state data and the fBIRN phantom, we used Weisskoff plots and guidelines of the fBIRN research group including average tSNR.[76-80] For anatomic quality (T1/T2 weighted), we incorporated metrics [62], including geometry accuracy, high contrast spatial resolution, the accuracy of slice thickness and position, image intensity homogeneity, and low-contrast object detectability. The QA/QC procedure was used to establish compatibility of data from different sites and long-term reproducibility of the results at each location.

### Data Collection, Data Transfer and Participant Data Acceptance

Data were acquired from multiple sites from two different scanner platforms, which are likely to produce different sample means as well as different variances. [59] Imaging data transfer from the study sites was performed through a secure virtual personal network (VPN) login into our network at Pediatric Imaging Research Center at Children’s Hospital of Pittsburgh contained within the University of Pittsburgh secure firewall. File transfer used an encrypted secured file transfer protocol (SFTP), with user authentication ensuring only approved users can connect to our portal. The upload portal was developed at the University of Pittsburgh, built on the XNAT framework. XNAT is an open-source informatics platform developed at Washington University specifically for high throughput management and sharing imaging data, including connectivity data. [66,81-83] This platform is highly extensible and contains a robust network security foundation. The portal has study specific user management, which allows the site-specific users to input de-identified participant and protocol information and images in native DICOM format, to enforce uniformity across sites. Furthermore, the XNAT framework provides for the incorporation of advanced processing pipelines, which allows each study site to perform data integrity checks and quality assurance before sharing the data. Long-term data storage was provided by servers at the University of Pittsburgh running a dedicated study specific PostgreSQL database and built-in parity against data loss. The PHN clinical data from the SVR studies in the form of SAS datasets (export files) were transferred from the PHN Data Coordinating Center for analysis with the imaging findings using a secure FTP site.

### Measures of Neurodevelopmental and Psychosocial Functioning

Standardized psychological assessments were performed by a psychologist and/or a supervised psychometrician at each site (Table 1). These measures were obtained in the SVR participants as part of the parent study [48] (Figure 3) and were explicitly included in our protocol for assessing the healthy controls. Comprehensive assessment of all domains of function including intellectual, language, visual spatial/nonverbal, learning/memory, fine motor, attention/executive, and social skills (Table 1). Caregivers completed standardized parent report measures for attention/executive, social-emotional, behavior, adaptive, and quality of life (Table 1). The testing required approximately five hours. Breaks were provided for snacks/lunch as appropriate for the participant. Evaluations occurred at least six weeks after any hospitalization. The psychologist or psychometrician at each site was blinded to the shunt type at the time of the Norwood operation.

**Table 1.** Neurocognitive Battery for SVR III Subject and Healthy Controls

| Domain | Instruments and subtests | Completion time- child | Completion time- respondent |
| --- | --- | --- | --- |
| Intelligence | Wechsler Intelligence Scale for Children-V (WISC-V; original test kit)<br>Block design<br>Similarities<br>Matrix reasoning<br>Digit span forward<br>Backward<br>Sequencing<br>Coding<br>Vocabulary<br>Figure weights<br>Visual puzzles<br>Picture span<br>Symbol search | 65-80 min |  |
| Math | Wechsler Individual Achievement Tests III (WIAT-III)<br>Math problem solving<br>Numerical operations | 30 min |  |
| Reading | Wechsler Individual Achievement Tests III (WIAT-III)<br>Word reading<br>Reading comprehension<br>Pseudoword decoding | 32 min |  |
| Language | NEPSY-II<br>Comprehension of instructions<br>Oromotor sequence | 13 min |  |
| Executive Function and Attention | Delis-Kaplan Executive Function System (DKEFS)<br>Tower<br>Trail making (all 5 Trials)<br>Verbal fluency (Letter, Category, Switching) Behavior Rating Inventory of Executive Functioning (BRIEF)<br>Parent and teacher report<br>Conners' III ADHD Index<br>Parent and teacher report | 35 min | 15 min<br>8 min |
| Visual and Perceptual Skills | Beery Developmental Test of Visual-Motor Integration (VMI-6)<br>Beery VMI<br>Visual perception | 8 min |  |
| Fine Motor | Lafayette Grooved Pegboard (use administration instructions from manual record raw time in seconds) | 5 min |  |
| Memory | Wide Range Assessment of Memory and Learning (WRAML-2)<br>Story memory, Story memory recall, Story recognition; Design memory, Design recognition; Verbal learning, Verbal Learning recall, Verbal learning recognition; Picture memory, Picture memory recognition; Finger windows; Number letter; Verbal working memory; Symbolic working memory; Sentence memory | 11 min | 7 min |
| Social Skills | NEPSY-II<br>Theory of Mind and Affect Recognition Autism Spectrum Rating Scale (ASRS) Parent report |  |  |
| Behavior | Behavioral Assessment System for Children – Second Edition (BASC-2)<br>Parent and teacher report |  | 20 min |
| Quality of Life | PedsQL Generic and Cardiac Modules Parent and Child reports |  | 15 min |
| Adaptive Function | Adaptive Behavior Assessment System – Third Edition (ABAS-3)<br>Parent report |  | 20 min |
| Total Time Required |  | 199-214 min | 85 min |

**Figure 2:**
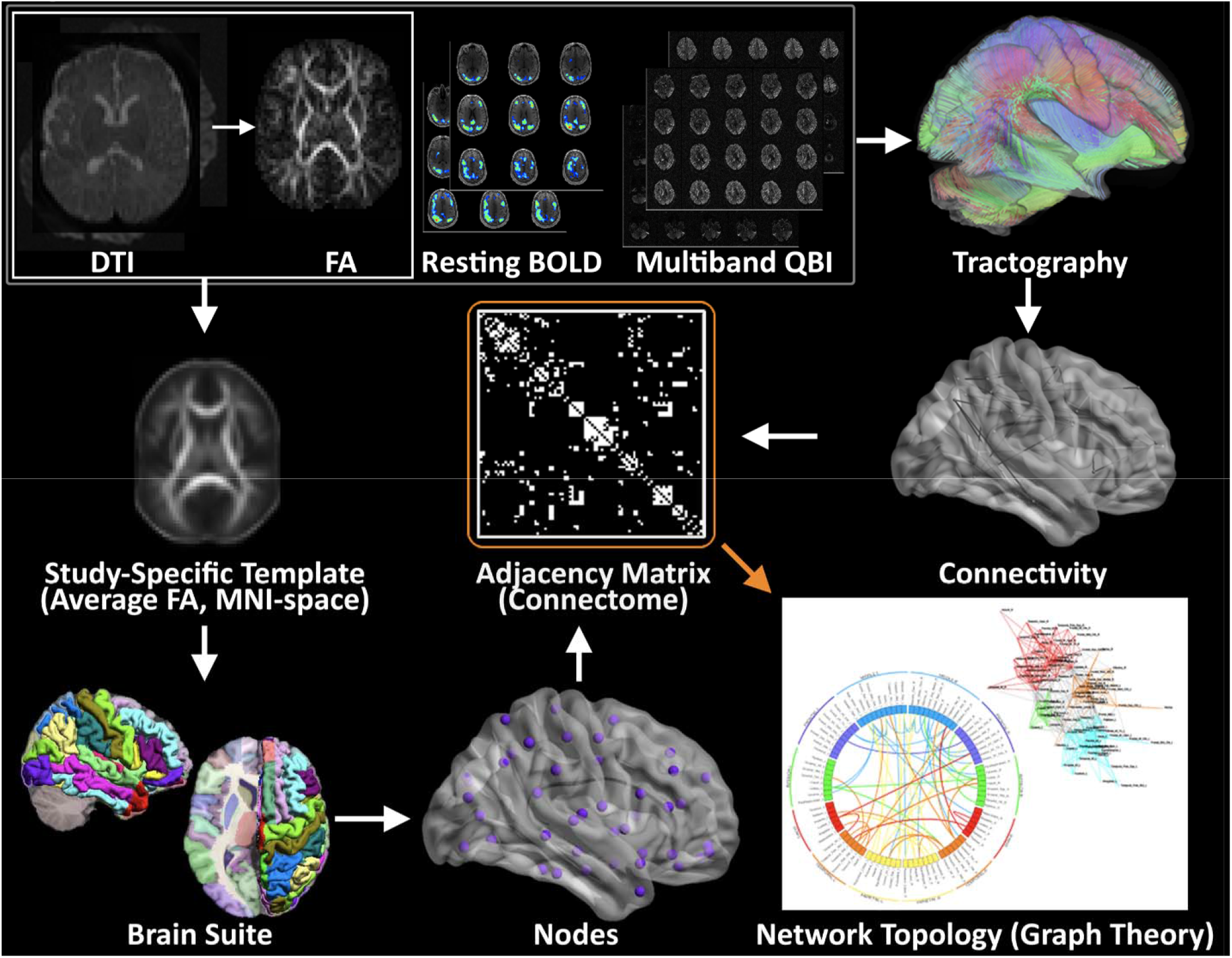
Methodology for Structural and Functional Network Topology Analysis. A flow diagram for the construction of WM structural and functional networks from DTI, resting BOLD and multi-band DTI including registration, segmentation, generation of WM fiber tracts using deterministic tractography, generation of adjacency matrix and nodes, and visualization of connectivity with spring-board and circle diagrams

**Figure 3:**
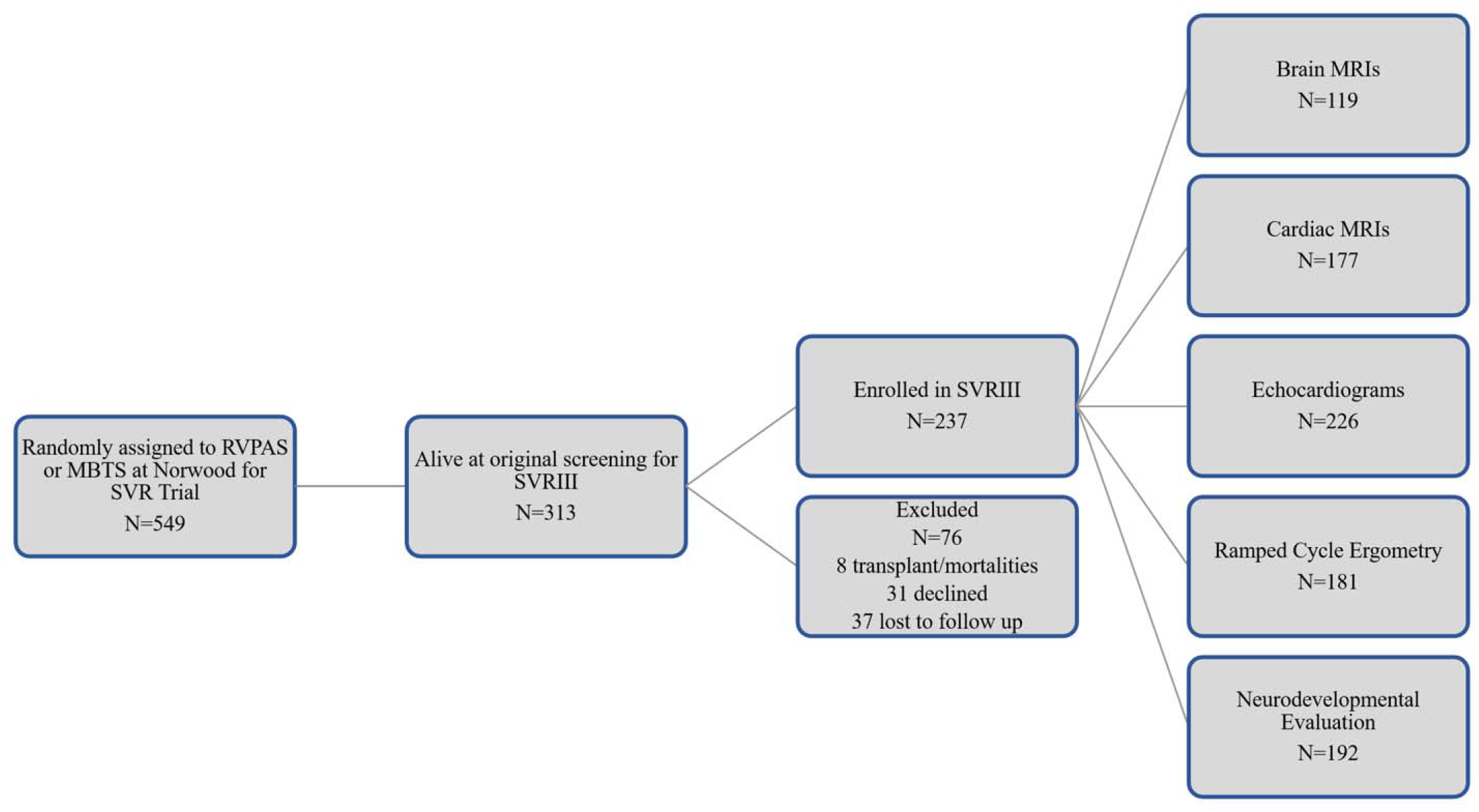
Original SVR Trial recruitment and SVRIII parent study enrollment

### Planned Imaging Post-processing

Standard 45 direction-DTI, Resting BOLD data, and MB-DTI (Figure 2) will be analyzed separately for the generation of graph data. *<u>DTI Pre-processing</u>*. All images will be corrected for motion, eddy current, and slice dropout artifacts using standard routines in FSL (FMRIB, Oxford UK). DTI data will be segmented into 90 cortical regions by applying the **Brain Suite Custom Atlas** [84-94] to produce cortical surface meshes and tissue classification maps.

#### Standard DTI

DTI metrics, including fractional anisotropy (FA), signal intensity without diffusion weighting (S0), and direction of the principal eigenvector, will be computed for each voxel. Deterministic tractography will be performed using in-house software written in Interactive Data language (IDL) (http://www.ittvis.com, Boulder, CO). Streamlines will be computed from each white matter voxel (determined as all voxels with FA > 0.25) in both directions. Stopping thresholds for the tractography will be turning angle > 45 degrees or FA < 0.25. *<u>MB-DTI</u>*. For the MB-DTI data, due to the large number of directions, the orientation distribution function (ODF) will be computed according to routines in DSI Studio.[95-97] Tractography will be performed according to routines in DSI Studio. The ODF allows for the detection of crossing fiber tracts within a voxel and thus allows a more accurate reconstruction of fiber tracts than is possible with standard DTI. A more accurate metric of anisotropy generalized fractional anisotropy (GFA) will also be computed. Streamlines will be computed from each white matter voxel (GFA > 0.25) and stopping thresholds will be turning angle > 45 degrees or GFA < 0.25.

#### Graph Construction DTI

Weighted graphs (estimate of connection strength between two regions) will be computed based on: 1) the total number of streamlines beginning and ending at two regions, and 2) the average FA or GFA value of all streamlines beginning and ending at two regions.

#### Resting BOLD

The Resting BOLD data will be processed through a robust motion detection and correction pipeline described by Powers et al[98,99], which involves volume censoring according to motion and intensity metrics, low-pass filtering, and regressing out of nuisance variables including motion parameters and global signal.

The resting BOLD data will also be parcellated using the Brain Suite atlas into 90 cortical regions. Correlation matrices will be constructed with each matrix element equal to the cross-correlation of the fMRI time series averaged in each of two regions.

#### Graph construction (Resting BOLD)

Binary unweighted graphs will be computed based on thresholding the correlation matrices at various values of cost ranging from 0.05 to 0.45.[100]

#### Graph analysis

Graph metrics (global efficiency, modularity, and transitivity) [101]will be computed via the C++ modules available from the Brain Connectivity Toolbox (BCT; Indiana University)-(For further details, see the BCT documentation at https://sites.google.com/site/bctnet/ and ref.[102]). Small-worldness, another graph metrics, will be computed using routines in IDL. For resting BOLD (Blood Oxygen Level Dependent), values of the graph theory metrics will be averaged over all values of cost.[103] At the nodal level, participation coefficient and clustering coefficient will be computed using the BCT. Spring load diagram and circle connectivity diagram will be used for visualization of relationships. [104,105]

### Planned Brain Connectome Outcome Measures

Primary outcome measure will include global graph measures derived from the 42-direction DTI data including global efficiency (primary) with modularity, transitivity and small-worldness measured secondarily; and also, sub-network (anatomically defined) and nodal level (degree, participation coefficient, nodal efficiency, clustering co-efficient) will be measured secondarily depending on the global metrics previously delineated. A similar approach will be applied to other imaging modalities collected (resting BOLD, HARDI, etc)

### Planned statistical analysis

Using a standard generalized linear model, we will compare global metrics (efficiency, transitivity, modularity and small-worldness) and sub-network/nodal metrics (clustering coefficient and participation coefficient) between the SVR III and control cases.

#### Analysis of indirect effects

To precisely determine the effect of single ventricle diagnosis on network topology (and, ultimately, on neurocognitive outcome), we will perform a statistical mediation analysis as we have previously defined. [106] In these analyses, single ventricle diagnosis status (verses control) will be the independent variable, neurocognitive outcome will be the dependent variable, and the graph metrics will be mediating variables (with the same covariates as the standard generalized linear model above). Bootstrapping (25,000 iterations, resampling with replacement) will be used to test for statistical significance, as the indirect effect (which is the product of two regression parameters) has a well-known non-normal distribution. Bias-corrected and accelerated confidence intervals,[107] shown to provide accurate control with optimal power for mediation analyses,[108] will be computed. Because many mediating variables will be tested, the false discovery rate (FDR) method[109] will be used to control for false positives at *q* < 0.05. Additionally, to assess the possible effects of perioperative variables, further analyses will be conducted on the cohort of SVR participants, with perioperative variables the independent variable, neurocognitive outcome the dependent variable, and graph metrics the mediating variables. These analyses will be performed on a global basis and only at a sub-network/nodal level as a *post hoc* analysis. Correlation of structure and function will be estimated with (1) visual correlations of nodal mapping, and (2) using AAL template anatomically-defined seed regions processing of resting state fMRI and DTI data.

#### Covariate Measures (Independent Risk Factors)

The unique nature of this large inception SVR III cohort presents the opportunity to examine the association between the clinical events that commonly occur in this population in relation to brain connectivity and neurodevelopment, as we did for the BCAS trial. Covariates that are available from the SVR trial and SVR follow-up studies (SVR II, III) include extensive pre-operative, peri-operative and annual follow-up measurements. We will prioritize those variables that are known to be associated with poor neurodevelopmental outcomes (Table 2).

**Table 2:** Examples of Longitudinal Clinical Risk Factors in the SVR Trial Dataset[48]

| Pre-stage I and Demographics | Stage I, II, III Hospitalization | Medication Use | Procedural History Data |
| --- | --- | --- | --- |
| • prenatal diagnosis<br>• genetic syndrome<br>• birth weight<br>• gestational age<br>• intubation<br>• socioeconomic status<br>• maternal education | • mechanical ventilation time<br>• cardiopulmonary bypass and deep hypothermic circulatory arrest/ regional cerebral perfusion times<br>• ICU/hospital length of stay<br>• delayed sternal closure<br>• use of extracorporeal membrane oxygenation (ECMO)<br>• unplanned surgical/catheter interventions | • treatment with heart failure medications (angiotensin-converting enzyme inhibitors)<br>• angiotensin receptor blockers, beta blockers<br>• treatment for pulmonary hypertension<br>• use of anticoagulation | • unanticipated interventions (catheterization or surgery, such as pacemaker implantation, stent implantation, Fontan revision)<br>• lack of Fontan completion |

#### Power analysis

We calculated required sample size for 80% power and two-sided α=0.05 to detect effect sizes estimated from the graph theory data on DTI in the BCAS (preliminary data) in relation to the global metrics for aims 1-3 using G*Power 3.1.3.

Effect sizes f^2^ were computed as (variance explained by effect)/(error variance), converted to effect size f for aim 1 as the desired input in G*Power 3.1.3. Effect sizes of f^2^ <0.15 (f<0.39) are considered to be small effects. <u>Aim 1:</u> The effect sizes for differences between SVR participants and controls are: (a) global efficiency: effect size f = 0.29, requiring a sample size of 97 subjects total for 0.8 power; (b) modularity: effect size f^2^ = 0.15 requires a sample size of 54 subjects total for 0.8 power; and (c) small worldness: an effect size of f = 0.34 will require a sample size of 69 subjects for 0.8 power. <u>Aim 2:</u> The effect size for the correlation between full-scale IQ and global efficiency (combined SVR/control group): is estimated to be f^2^ = 0.093, requiring a sample size of 87 subjects for 0.8 power. A similar given power was detected for other domains (visual spatial, memory, executive function) which were also tested in the BCAS study. <u>Aim 3:</u> The effect size for the correlation between total cooling time and global efficiency in the SVR cohort is estimated to be f^2^ = 0.096, requiring a sample size of 84 subjects for 0.8 power and 0.05. These calculations demonstrate that the required sample size to detect effect sizes similar to those observed in the BCAS with 80% power are all smaller than our initially targeted sample size of 140 SVR subjects and 100 referents; furthermore, the study is well-powered to detect small effect sizes for our study hypotheses. From this power analysis, we determined that a sample size of at least 100 SVR subjects with analyzable MRI data would be ideal. Given that we expected a certain portion of SVR participants to fail the imaging procedures (based on non-compliance and too much motion artifact), we decided to initially target approximately 140 total SVR participants, expecting 1/3 of data points to be unanalyzable. We also initially targeted 100 age-matched controls to be recruited from the same sites. Therefore, our initial planned targeted enrollment for the study was approximately 240 subjects (SVR+Controls).

### Initial Recruitment Challenges

In this ancillary study of the longitudinal SVRIII parent study, we aimed to add a brain imaging component in addition to the parent’s study multidisciplinary evaluation including the performance of a cardiac MRI study, echocardiogram, exercise testing, and neurocognitive evaluations over two full days (Figure 1). To avoid interference with the parent study, we performed the brain MRI component after the measures required for the parent study. However, while most participants completed the full program for the parent study, some participants and parents deemed the extra allotted time for the MRI too taxing, especially after the neurocognitive assessment. While incentives and extra travel accommodations were available to reschedule the brain MRI, some participants were lost to this ancillary study who were enrolled in the main SVR study.

To counteract participant fatigue and avoid losing participants, we devised a strong coordinator-driven explanation of the study, and direct benefits to the participants and indirect benefits to the community in general to increase interest. We also increased the number of meetings with direct key study personnel (study investigators e.g., doctors, coordinators, MRI specialists) to enhance successful participant recruitment for this ancillary study. We devised a bi-monthly call between coordinators of all sites to discuss challenges and help each other identify various successful recruitment strategies. These twice-monthly multi-site coordinator calls were extremely well-attended and allowed communication between sites regarding recruitment difficulties, addressing MRI safety and screening issues, and reviewing case report forms and protocol revisions. Strategies to recruit healthy controls were expanded at multiple sites to include the use of clinical translational research registries, pediatric clinics seeing healthy patients, and community sites (schools, churches, and special neighborhoods). Of note, ADHD is more prevalent in the HLHS population, and our neuroimaging protocol was designed to maximize data acquisition but minimize scanner time. We also increased the number of enrolling sites to assist with the enrollment of both SVR and control subjects.

### Initial Technical Challenges

Some of the initial technical challenges of the study were related to imaging acquisition, imaging harmonization, and data transfer/storage. With regard to image acquisition, there were challenges to the acquisition of multi-band (MB) BOLD and HARDI scans (Fig. 7). Many of the errors were operator errors. Both MB scans require a reference scan to be acquired prior to running the main body of sequences for purposes of reconstruction. Hence, running the main body of sequences without first running the reference scan was a common mistake. A related mistake was copying scan parameters from BOLD images or using the BOLD reference image for HARDI sequences, which have different image and slice resolutions, resulting in lower resolution HARDI images and an unusable data set. Another common mistake was prescribing an inadequately-sized shim box resulting in distorted HARDI images. Less frequent, but still encountered was failure to turn on coil elements leading to images with low SNR. Lastly, in a few cases, fat saturation bands, used to estimate multiband coverage by some sites, were left on, leading to images with a swath of HARDI image slices with suppressed signal. These errors were documented in a technical manual and information was disseminated to all sites to prevent these errors from recurring. In addition, frequent site visits were conducted to provide education about these issues.

**Figure 4:**
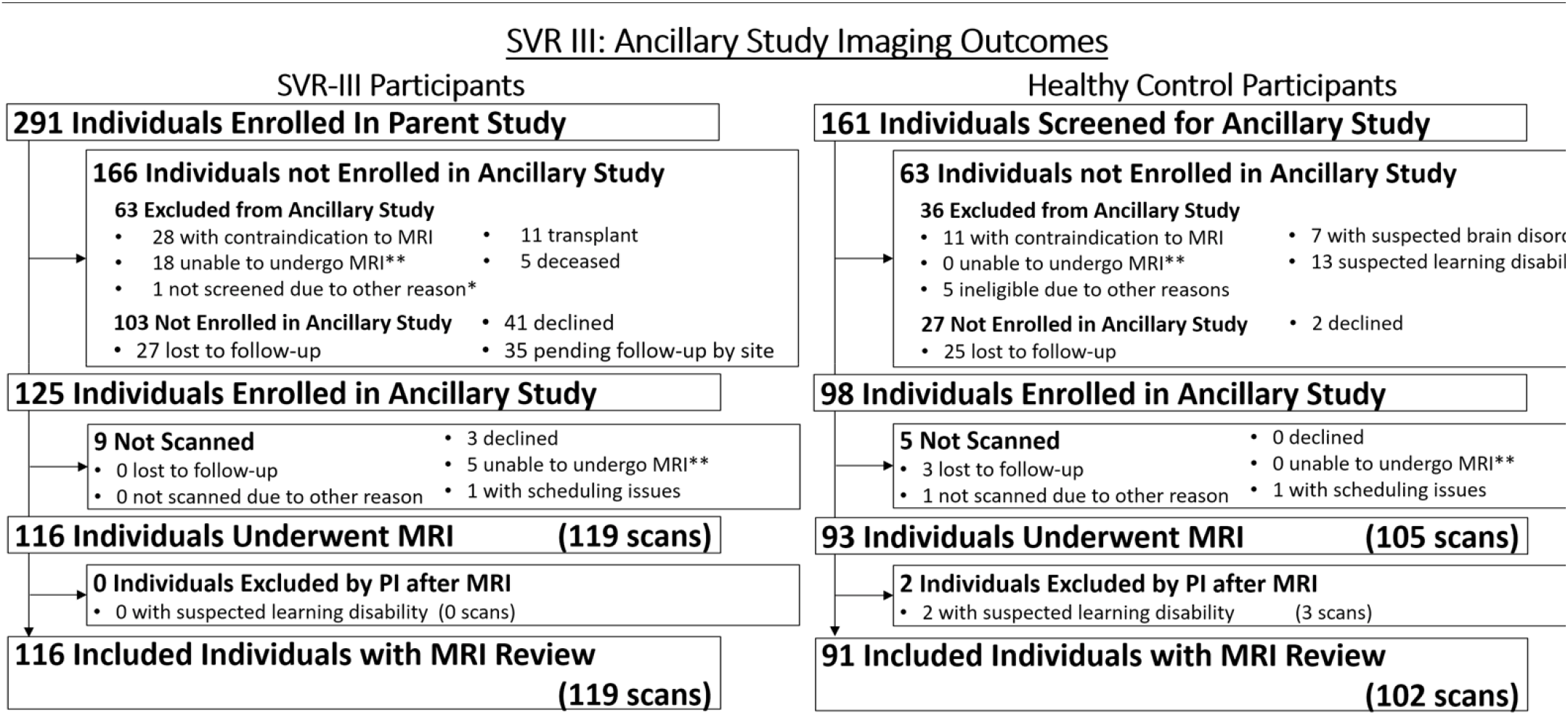
SVRIII Brain Connectome: Enrollment and Collections of Neuroimaging Studies for SVRIII Participants and Healthy Controls

**Figure 5:**
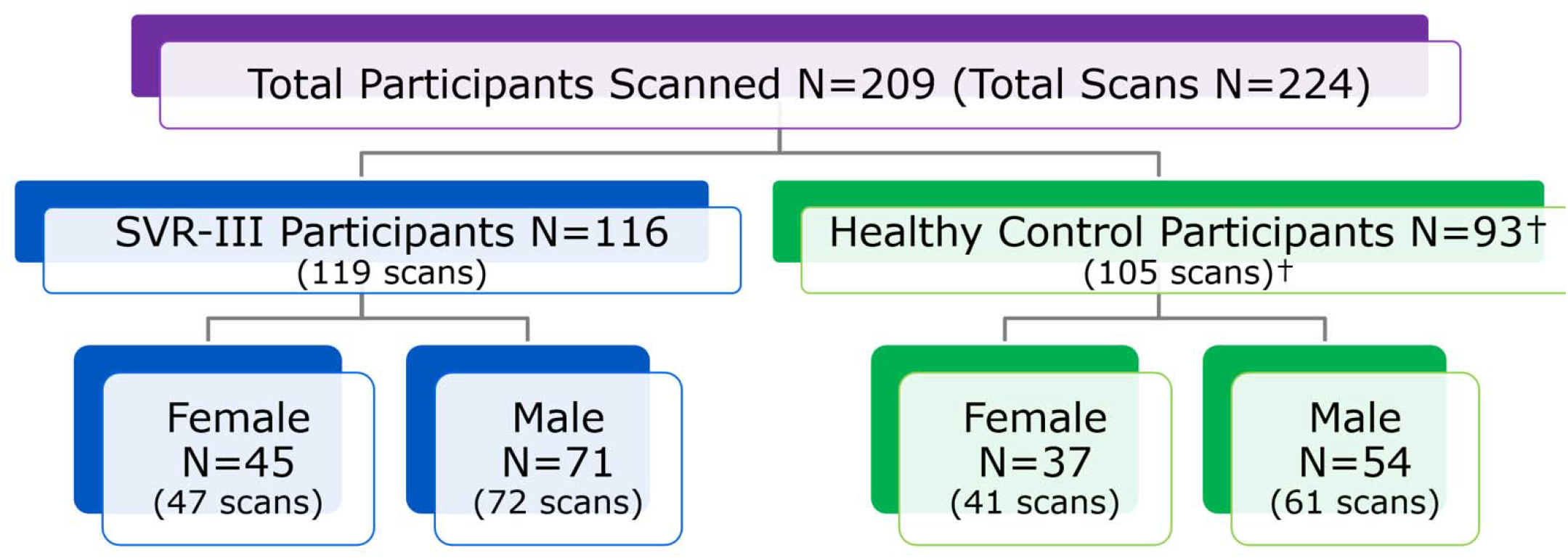
SVR III Brain Connectome Study: Diagnostic group and postnatal sex of enrolled participants.

**Figure 6:**
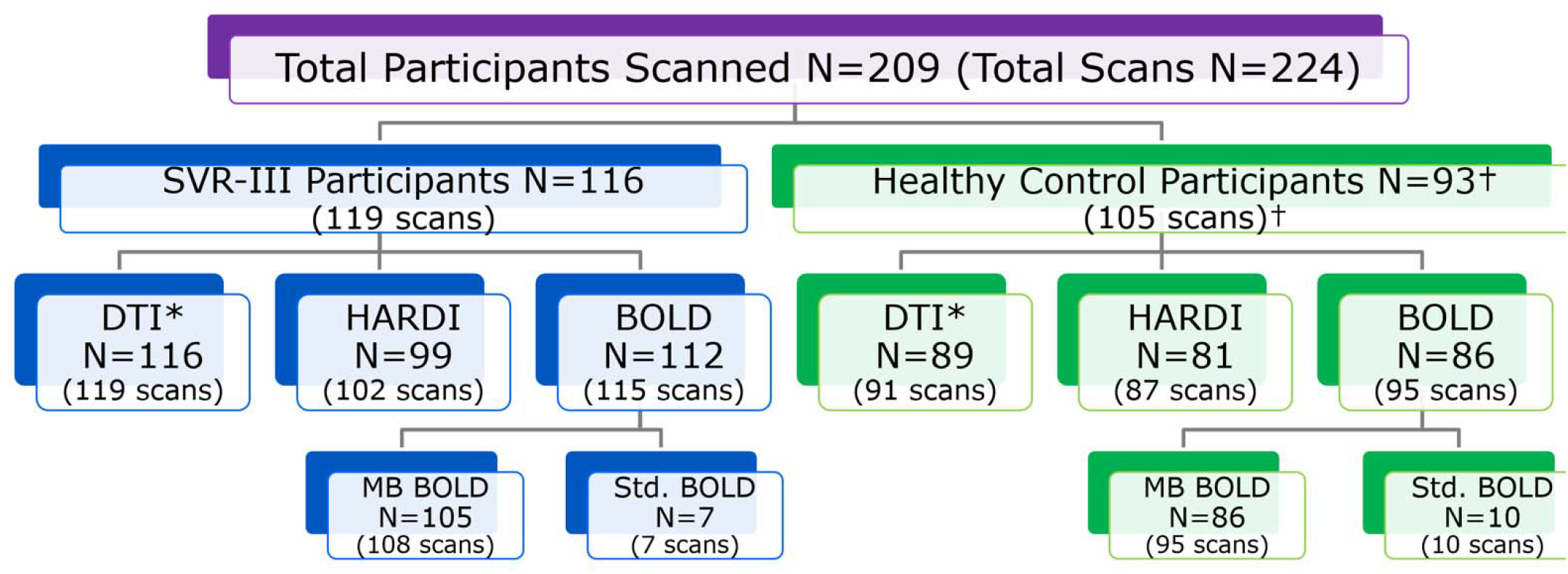
SVRIII Brain Connectome Study: Breakdown of imaging studies collected (DTI,HARDI, Resting BOLD).

**Figure 7:**
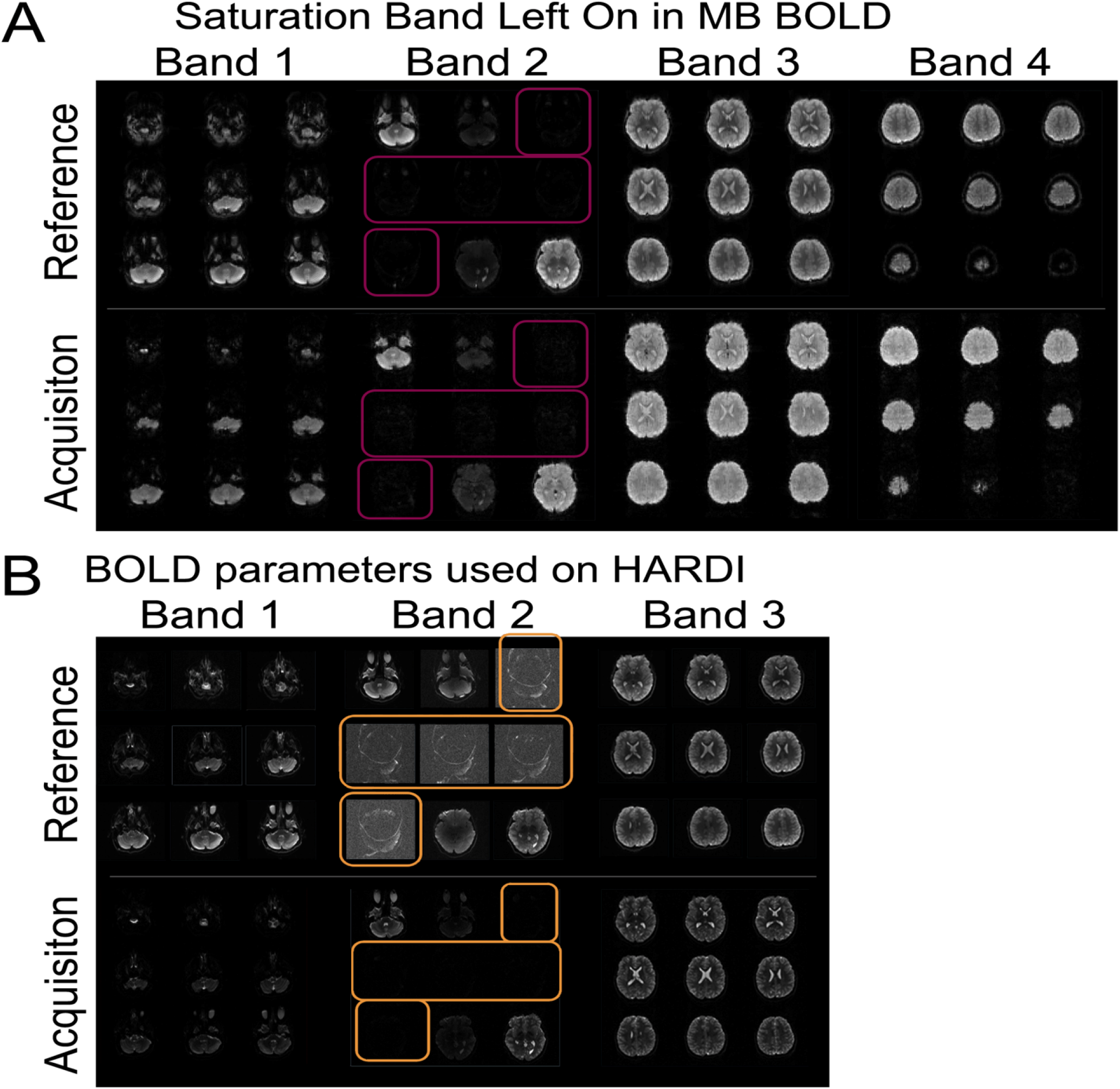
Multi-band reference scan error example with A: Saturation band left on the BOLD imaging and B: ultimately propagated to the DTI/HARDI prescription and acquisition in error. These errors underscore the need for a technical manual, ongoing QA of data, and technological training.

With regard to imaging harmonization, given the anticipated scanner variance from using two different vendor scanners, QA/QC procedures include a complex multi-step harmonization process that involves; (1) development and surveillance of a standardized neuroimaging protocol across sites; (2) prospective on-going synthetic and human phantom studies to provide cross-calibration across scanner; and (3) retrospective or statistical harmonization of human subject neuroimaging data, knowing that inter-scanner variation will still occur despite implementation of (1) and (2). To address some of these issues, we increased the number of both synthetic and human phantom data (a total of five human phantoms and two synthetic phantoms scanned three times) (Figure 8). We specifically introduced the use of a HARDI phantom given the data that were being collected in the study, knowing that diffusion imaging tends to have the greatest inter-scanner variance even after protocol matching. We currently have a database of greater than 100 synthetic and single human phantom HARDI diffusion tensor data on PHN 3T MRI scanners (Siemens /Phillips vendors). We have recently developed a new pipeline that incorporates both synthetic phantom and human individual subject-specific template/tract generation via a semi-automated approach.

**Figure 8:**
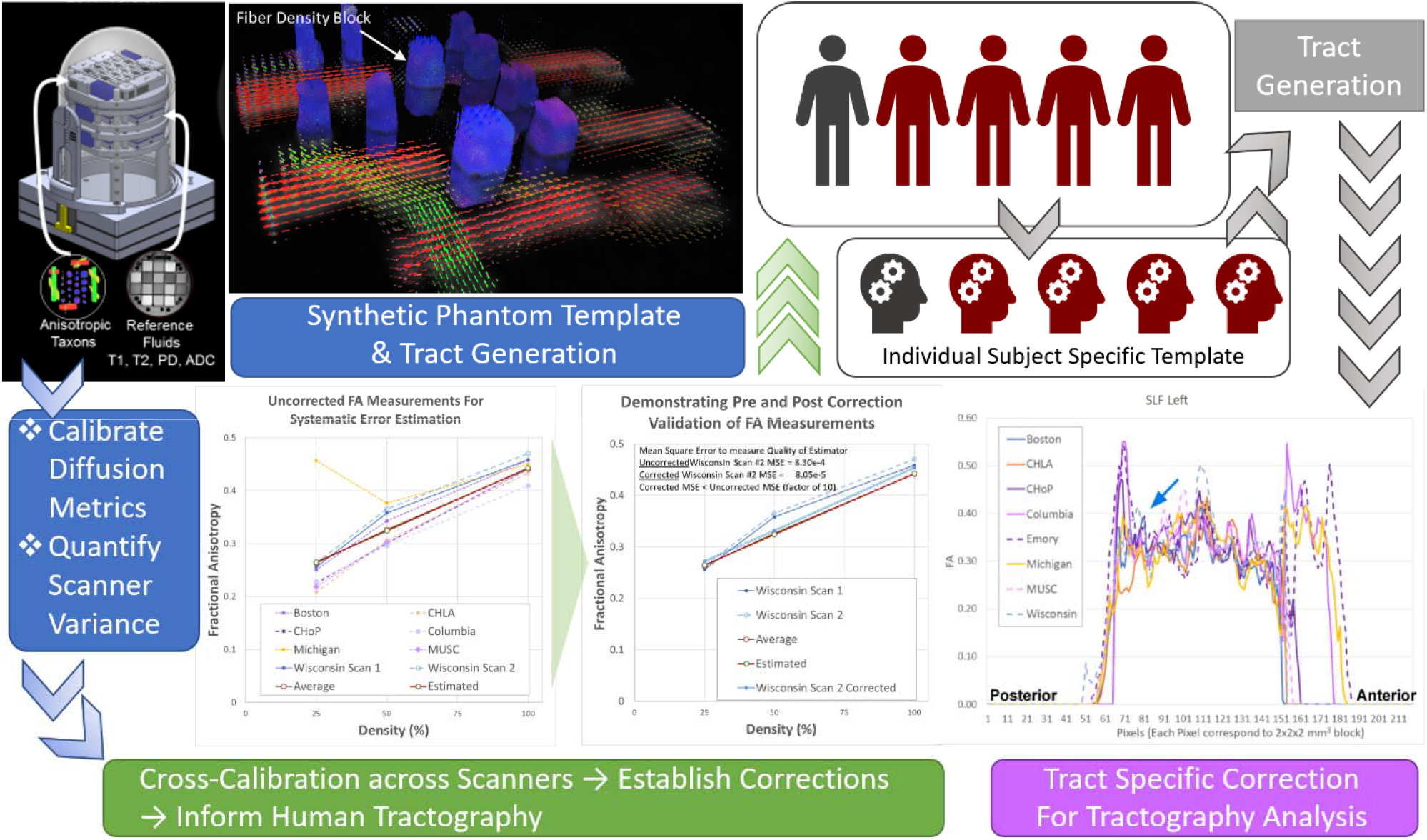
Harmonization Design: We used synthetic and human phantom and semi-automated pipeline to generate tract-specific information to estimate interscanner variance and then applied to the in vivo analysis; Bottom row-left: FA measurements of taxon blocks from seven PHN sites. The results show a reduced variability in the calibrated FA in the corrected scan (blue double line) compared to the uncorrected scan (blue dotted line). Bottom-right: Along-tract FA analysis of an individual human phantom across multiple sites. The tract shown is the left SLF showing portions of tract that are more sensitive to scanner variability (blue arrows).

This dataset will allow for future analyses that can directly estimate the effect of scanner type. For diffusion MRI, our synthetic phantom has been developed which simulates various axonal configurations including varying density, fiber crossing, etc., constructed from textile “taxons” of similar diameter to actual axons. By scanning the phantom repeatedly across sites, estimates may be obtained for within-site and across-site variability, and these estimates may be incorporated into the final analysis. However, this approach is limited by the type of diffusion metrics that can be estimated using the phantom, such as voxelwise fractional anisotropy or mean diffusivity and is unavailable if more advanced diffusion metrics (e.g. along-tract FA/MD values derived from tractography, estimates of myelin water fraction, graph analysis parameters, etc.) are desired that cannot be computed from the phantom. For these more advanced metrics, we will be able to utilize our human phantoms, which are scanned repeatedly at each site. Again, these data will yield estimates of within- and across-site variability for each desired parameter, which can be incorporated into the final analysis.

With regard to image transfer, some of the sites were within secure hospital firewalls, while other sites were within pure research imaging environments. To address the variability in firewalls and ability to transfer de-identified neuroimaging data, we set up our study database using the open-source neuroimaging specific framework XNAT. We then customized a secure, externally accessible portal on our University of Pittsburgh domain with standardized forms for demographics and data entry. Our protocol provided two options for each site to upload imaging data: A) directly to our XNAT database, accessible via the web portal and B) via an sFTP transfer into a local server behind our firewall. Option A was the preferred method, as images were directly archived into our database and linked to the appropriate subject data. We required sites to anonymize the studies prior to uploading (regardless of upload method), and our

XNAT server performed a secondary safety-net anonymization prior to archiving in case any remaining personal health information were present in the DICOM headers. At time of development, the upload portal required a Java plugin to upload DICOM studies. Due to varying degrees of internet security measures at each site, some of the clinical sites were restricted from using a Java plugin. In these instances, if IT support was not provided to implement a white-list exception to this plugin, sites were unable to upload the images via this method. Subject forms (including consent, demographics, and behavioral testing) were still entered here, however, as no plugin was necessary. Thus, an option B was presented as an alternative to image transfer. Via option B, DICOM studies were uploaded into our local server, and our own database manager archived it in the XNAT database. This method was used solely on a need-basis, as it required an additional step for our coordinating site, and sites were required to receive University of Pittsburgh guest accounts with 2-factor authentication (2FA), which can be a technical challenge for some users. Finally, if a site was unable to use methods A or B due to technical, security, or personnel difficulties, we requested that physical copies of the anonymized images be sent via secure courier to our facility for archival by our database manager. Updates to our technical protocol and frequent site visits to troubleshoot these image transfer issues helped with developing a robust image transfer process.

### Timeline and Impact of COVID-19 Pandemic

The first participant in the SVRIII Brain Connectome study was enrolled on April 6, 2016, nine months after the first study participant was enrolled in the parent SVR III study. Planned enrollment was predicted to last until approximately April 2020 (4 years).

We prolonged the extension of this ancillary study after the parent study finished enrolling in September 2019, to increase study participation up until January 2022. Importantly, the COVID-19 pandemic prevented the completion of in-person evaluations, more common over the summer, between March 2020 and August 2020 at most sites (Figures 3-6). The SVRIII Brain Connectome study, which finished enrolling in January of 2022, enrolled 125 SVRIII participants (116 analyzable scans) and 93 control participants (89 analyzable scans). From our power analysis (see power analysis section), we determined that a sample size of at least 100 analyzable SVR subjects would be ideal. As such, the final enrollment number of n=218 subjects did achieve our goal.

## Discussion

The SVRIII Brain Connectome study will be the first to validate connectome neuroimaging biomarkers to prognosticate outcomes in early adolescent HLHS patients. It leverages a rich longitudinal dataset of HLHS patients collected as part of the parent study. The SVRIII Brain Connectome study will not only help to elucidate the impact of complex CHD on brain development, but also the manner in which a developing neural architecture— connectome—gives rise to cognitive-behavioral phenotypes in the SVRIII longitudinal cohort. The data generated will allow us to determine whether brain network topology can serve as a biomarker for specific behavioral and neuropsychiatric phenotypic deficits and whether peri-operative and patient factors are associated with the development of specific brain network topology. The results of these studies will provide the basis for future predictive modeling and ultimately, targeted interventions to improve neurodevelopmental outcomes of HLHS patients.

Our study overcame several obstacles, as described in detail above. Initial lagging recruitment was successfully addressed by multiple approaches, including addition of recruiting sites and frequent multi-site coordinator calls to maintain enthusiasm and brainstorm about recruiting techniques. Technical challenges that emerged early in the study were related to neuroimaging acquisition, harmonization and transfer. We responded to these challenges with modification of the study protocol and frequent site visits which involved traveling human and synthetic phantoms. A specific challenge of multi-site MRI studies is harmonization of MRI data across different sites and scanner platforms. Between different MRI vendors (e.g. Philips, Siemens, GE), the same apparent MRI pulse sequence (e.g. identical parameters are entered at the console) is quite often variable in practice due to RF pulse shape, diffusion-encoding gradient (e.g. amplitudes and duration can be vendor distinct even for the identical “b-value” entered at the console), bandwidth in slice-select direction. For the same MRI vendor but a different scanner platform (e.g. Siemens Skyra vs. Siemens Prisma), the actual pulse sequence is often different due to different hardware specifications (e.g. maximum gradient strength, slew rate, RF power, etc.) between the two platforms, as the actual pulse sequence run is optimized dependent on the available hardware.

We have recently developed multiple retrospective harmonization approaches that can eventually be applied to this dataset, if needed. For example, if the number of subjects per site is adequate, and the SVR III participant/control ratio is approximately equal across sites, data may be analyzed separately for each site and the final result combined afterwards, with negligible loss of statistical power. We have previously used this approach in a two-site study comparing DTI analyses (graph analysis and voxelwise) between neonates with CHD and healthy controls[47]. Data were analyzed separately to estimate the between-subject variance separately for each platform, and for the site-specific estimates were then combined to perform an aggregate analysis (with scanner platform incorporated as a nuisance parameter. Another retrospective neuroimaging harmonization approach is the empirical Bayes method (COMBAT). If the total sample size is sufficiently large, it is possible to estimate a (site-independent) prior distribution from the data itself. Site-specific correction factors are then able to be estimated and applied to the data. The final analysis is performed using the combined, harmonized data. We have recently successfully applied this COMBAT technique to a four-center DTI dataset of 763 neonates with congenital heart disease.[110]

## Conclusions

The SVR III Brain Connectome study leverages the PHN SVR III study, adding brain MRI and the inclusion of healthy controls to undergo neurocognitive evaluation and brain MRI to understand not only the basis of important brain-behavior phenotypes that are present in single ventricle patients, but also the modifiable and non-modifiable longitudinal risk factors that predict these relationships. Because this was a multicenter brain imaging study, harmonization was prioritized with initial protocol matching, use of human/synthetic phantoms and development of novel retrospective neuroimaging techniques. Through the conduct of this study, we learned about some of the challenges and solutions related to adding neuroimaging ancillary procedures to a parent study design. Analyses will elucidate not only the impact of single ventricle heart disease on brain development, but also the relationships between the human connectome and cognitive-behavioral phenotypes.

## Data Availability

All data produced in the present study are available upon reasonable request to the authors

## Author Contribution

Conceptualization, A.P., V.S., and R.C.; Methodology, A.P., V.S., and R.C.; Software, V.S. and V.L.; Validation, A.P., V.S., R.C. and V.L.; Formal Analysis, A.P., V.S., R.C., V.L., and J.W.; Investigation, J.W.; Resources, A.P.; Data Curation, V.S., R.C., J.W.; Writing – Original Draft Preparation, V.S., R.C., V.L., J.W., A.S., T.C, H.P., J.B., A.V., D.Q., N.K., P.G., B.G., P.L., M.M., L.S., D.B., D.I., S.O., T.M., J.D., K.H., A.A., M.R., J.C., W.M., N.G., W.G., C.G., J.N., and A.P. ; Writing – Review & Editing, V.S., R.C., V.L., J.W., A.S., T.C, H.P., J.B., A.V., D.Q., N.K., P.G., B.G., P.L., M.M., L.S., D.B., D.I., S.O., T.M., J.D., K.H., A.A., M.R., J.C., W.M., N.G., W.G., C.G., J.N., and A.P.; Visualization, A.P., and V.S..; Supervision, A.P..; Project Administration, A.P.; Funding Acquisition, A.P.

## Institutional Review Board Statement

IRB approval was obtained at each participating site and written consent obtained from parents prior to participation in SVRIII Brain Connectome. Additional oversight is provided by an NIH-appointed Data and Safety Monitoring Board and medical monitor. Parents and/or guardians are consented, and children assented (when possible) by the site PI or research coordinator for enrollment.

## Informed Consent Statement

Informed consent was obtained from all subjects involved in the study.

## Data Availability Statement

No new data were created or analyzed in this protocol/methods manuscript. Data sharing does not apply to this article.

## Acknowledgments

We acknowledge the hard work of the Pediatric Heart Network coordinators including Katherine Afton, Carolyn Dunbar-Masterson, Lisa Jean Buckley, Kathy Lupton, Michelle Otto, Regina Cole, Michelle Hamstra, Madison Rudow, Katrina Golub, Chanel Rojas, Mingfen Xu, Kalyan Chundru

