## Supplemental Table 1 for "Single Ventricle Reconstruction III: Brain Connectome and Neurodevelopmental Outcomes: Design, Recruitment, and Technical Challenges of a Multicenter, Observational Neuroimaging Study"

**Supplemental Table 1: SVRIII Brain Connectome Protocol: Harmonized Parameters for Siemens and Phillips 3T MRI**

|  |  |  | **Matrix** | **slices** | **FOV** | **% FOV phase** | **Resolution (mm)** | **TR (ms)** | **TE (ms)** | **TI (ms)** | **Flip Angle (deg)** | **Parallel Imaging** | **Multi-band Acceleration** | **Phase partial Fourier** | **Diffusions Directions** | **b-values** | **Acquisition Time** |
| --- | --- | --- | --- | --- | --- | --- | --- | --- | --- | --- | --- | --- | --- | --- | --- | --- | --- |
| Diffusion Imaging | Siemens | DTI | 128 x 128 | 60 | 256 x 256 | 87.50% | 2.0 x 2.0 x 2.0 | 8600 | 79 | N/A | 90 | 2x | Off | 6/8 | 42 | 1000 | 7:27 |
|  |  | HARDI | 100 x 100 | 60 | 240 x 240 | 100% | 2.4 x 2.4 x 2.4 | 3300/3600 | 110/131 | N/A | 90 | Off | 3 | 6/8 | 64 x 4 | 3000, 5000 | 3:54 x 2, 4:16 x 2 |
|  | Philips | DTI | 128 x 128 | 60 | 256 x 256 | 87.50% | 2.0 x 2.0 x 2.0 | 6640 | 75 | N/A | 90 | 2x | Off | 6/8 | 42 | 1000 | 7:31 |
|  |  | HARDI | 100 x 100 | 60 | 240 x 240 | 100% | 2.4 x 2.4 x 2.4 | 3033/3266 | 105/116 | N/A | 90 | Off | 3 | 6/8 | 64 x 4 | 2500, 4000 | 4:14 x 2, 4:46 x 2 |
| Resting BOLD functional MRI | Siemens | rs-fMRI | 64 x 64 | 36 | 256 | 100% | 4.0 x 4.0 x 4.0 | 650 | 32 | N/A | 50 | Off | 4 | 6/8 | N/A | N/A | 5:09 x 2 |
|  | Philips | rs-fMRI | 64 x 64 | 36 | 256 | 100% | 4.0 x 4.0 x 4.0 | 800 | 32 | N/A | 90 | Off | 4 | 6/8 | N/A | N/A | 5:07 x 2 |
| 3D T1&T2 | Siemens | T1 | 256 x 256 | 160 | 256 x 256 | 87.50% | 1.0 x 1.0 x 1.0 | 2400 | 3.16 | 1200 | 8 | 2x | Off | Off | N/A | N/A | 6:18 |
|  |  | T2 | 256 x 256 | 160 | 256 x 256 | 100% | 1.0 x 1.0 x 1.0 | 3200 | 411 | N/A | Variable | 2x | Off | Off | N/A | N/A | 3:30 |
|  | Philips | T1 | 256 x 256 | 160 | 256 x 256 | 87.50% | 1.0 x 1.0 x 1.0 | 1870 | 3.3 | 900 | 8 | 2x | Off | Off | N/A | N/A | 4:02 |
|  |  | T2 | 256 x 256 | 160 | 256 x 256 | 100% | 1.0 x 1.0 x 1.0 | 2500 | 253 | N/A | Variable | 2x | Off | Off | N/A | N/A | 4:28 |
